# Visual overactivation during metaphor processing: A novel mechanistic account of concretism in schizophrenia

**DOI:** 10.64898/2026.08.07.26359933

**Authors:** Valentina Bambini, Federico Frau, Chiara Pompei, Luca Bischetti, Veronica Mangiaterra, Ginevra Martinelli, Chiara Battaglini, Lorenzo Vita, Giulia Agostoni, Margherita Bechi, Mariachiara Buonocore, Jacopo Sapienza, Francesca Martini, Marco Spangaro, Federica Cocchi, Roberto Cavallaro, Marta Bosia

**Author notes:** **Corresponding Author:** Federico Frau, Laboratory of Neurolinguistics and Experimental Pragmatics (NEPLab), Department of Humanities and Life Sciences, University School for Advanced Studies IUSS, Pavia, Piazza della Vittoria 15, 27100 Pavia, Italy. equally contributed as co-first authors (Valentina Bambini, Federico Frau).

## Abstract

Individuals with schizophrenia show well-documented impairment in metaphor comprehension, often exhibiting a bias toward concrete, literal interpretations. While this tendency has traditionally been linked to psychopathological and cognitive factors, the contribution of perceptual processes remains underexplored. Here, we tested the hypothesis that figurative language impairment reflects altered perceptual processing, whereby the visual representations evoked by metaphors remain abnormally active and hinder abstraction. A sample of 143 individuals with schizophrenia and healthy controls was administered a novel paradigm where metaphors (e.g., *Wisdom is a flashlight*) served as primes for target words related to the metaphor vehicle based on visual (e.g., *microphone*), action (e.g., *remote*), or semantic features (e.g., *lamp*). While in healthy participants metaphors activated semantically associated words, individuals with schizophrenia showed sustained visual priming, emerging 1000 ms after metaphor presentation and persisting up to 1400 ms, with both groups showing reverse priming for action targets. Critically, greater visual priming predicted lower metaphor comprehension in patients, whereas greater semantic priming was correlated with better metaphor skills in controls. These results suggest that visual-perceptual representations are not only overactivated in patients compared to controls during metaphor processing but may also interfere with figurative comprehension. We argue that concretism arises from an imbalance between bottom-up sensory signals and top-down contextual priors, leading to the persistence of the visual representations and impaired abstraction. More broadly, these results support multimodal and predictive accounts of metaphor processing and point to altered perceptual dynamics as a previously unappreciated mechanism contributing to pragmatic impairment in schizophrenia.

## 1. Introduction

Classic psychopathology identifies impaired comprehension of figurative language, i.e., expressions whose meaning goes beyond the literal sense of words, as in the case of metaphors, among the core manifestations of formal thought disorder in schizophrenia (Bleuler, 1911; Goldstein, 1959; Harrow, 1974). Individuals with schizophrenia frequently exhibit a bias toward literal, concrete interpretation of metaphor (Deamer et al., 2019; Kircher et al., 2007; Mashal et al., 2013), as well as related phenomena such as idiom (Schettino et al., 2010; Sela et al., 2015; Titone et al., 2002) and proverbs (Brüne & Bodenstein, 2005; Felsenheimer & Rapp, 2023). While clinically this pattern is captured by the construct of *concretism*, defined as reduced capacity for abstract thought that goes beyond immediately available physical stimuli (Harrow, 1974), more recent accounts also frame it within the broader pragmatic disorder of schizophrenia (Bambini et al., 2016, 2020), i.e., as part of a more general difficulty in using language in a contextually appropriate manner.

When it comes to understanding the causes of figurative language impairment in schizophrenia, studies have highlighted a number of factors. From the psychopathological point of view, difficulties in understanding metaphors have typically been associated with negative symptomatology (Bambini et al., 2020; Kircher et al., 2014), but increasingly also with the positive and disorganization dimensions (Bischetti et al., 2026). Also, several studies have pointed to the role of cognitive deficits (Frau et al., 2024), in particular executive impairment, in handling and suppressing literal senses (Li et al., 2017; Schettino et al., 2010), as well as theory of mind difficulties in inferring intended meanings (Brüne & Bodenstein, 2005; Frau, Bosia, et al., 2025; Langdon et al., 2002; Parola et al., 2020). At the brain level, altered patterns of recruitment of the fronto-temporal cortex have been associated with metaphor impairment in schizophrenia (Kircher et al., 2007).

One factor that these accounts do not fully capture is the contribution of perceptual systems to figurative meaning derivation. Converging evidence from neuropragmatics (Bischetti, Frau, et al., 2024; Tomasello, 2023) indicates that understanding non-literal meanings recruits a distributed network of brain regions that includes prefrontal and temporo-parietal regions distributed bilaterally (Bambini et al., 2011; Rapp et al., 2012; Reyes-Aguilar et al., 2018), together with activations within the sensory-motor systems (Desai et al., 2011). Within this framework, metaphor is said to be supported by embodied simulation, whereby perceptual and motor representations associated with the literal meaning are reactivated and integrated into higher-level conceptual structures (Cuccio, 2022). Among the sensory-motor aspects evoked by metaphor, visual processing is particularly relevant, as it captures the longstanding intuition that metaphorical language is vivid and capable of ‘setting things before the eyes’ (Katz, 1998). The activation of visual, image-based representations in metaphor is supported by behavioral and neurophysiological studies (Al-Azary & Katz, 2021; Frau, Tomasello, et al., 2026; Khatin-Zadeh & Banaruee, 2026), which suggest that such perceptual activations are involved in the inferential pathway that leads to figurative interpretation (Carston, 2018; Gibbs & Matlock, 2008). Applied to the context of impairment, there is evidence that the disruption of sensory-motor systems might underlie figurative language difficulties in neurological patients (Fernandino et al., 2013; Frau, Diamanti, et al., 2026). Yet, whether altered sensory-motor processes contribute to concretism in schizophrenia remains largely unexplored.

In this study, we tackled the novel hypothesis that figurative language impairment in schizophrenia may be partly related to altered sensory-motor processing. In particular, we propose that the visual representations evoked by metaphors may be overactivated, leading patients to retain the physical meaning of concepts and thereby hindering abstraction. This interpretation is broadly consistent with predictive-processing accounts, according to which schizophrenia involves abnormalities in the integration of bottom-up sensory signals and top-down priors (Corlett et al., 2019; Sterzer et al., 2018; Weilnhammer et al., 2020). Extended to the case of metaphor comprehension, we reason that such an imbalance would result in perceptual representations associated with the literal meaning persisting more strongly than contextual information supporting the intended figurative interpretation. Hence, predictive processing provides a possible computational framework for the present hypothesis, which we test here through its behavioral consequences. More generally, we propose that visual overactivation during metaphor processing constitutes a novel mechanistic account of concretism in schizophrenia, linking language impairment to alterations in perceptual processing and offering a bridge between sensory-motor theories of meaning and computational accounts of psychosis.

Several strands of evidence support such a hypothesis. First, recent findings showed that patients with schizophrenia tend to use more concrete words compared to controls when asked to explain the meaning of figurative expressions (Bambini et al., 2025). For instance, patients may explain a metaphor such as *some memories are thorns* by saying that *some memories itch*, whereas controls rely on more abstract words (e.g., *some memories are sorrowful*). The higher concreteness at the semantic level might be linked to enhanced activation of the perceptual properties of concepts. Second, atypical engagement of the visual representational system has been reported in schizophrenia, affecting both perceptual processing and higher-order forms of imagery (Dijkstra et al., 2025). Disturbances encompass difficulties in integrating sensory signals into coherent object representations and in exploiting contextual information to guide perception (Silverstein & Keane, 2011), up to experiencing more vivid and intrusive mental images than healthy controls (Oertel et al., 2009; Sack, 2005; Stephan-Otto et al., 2017). These findings strengthen the possibility that altered visual processing may cause the persistence of physical properties of concepts, thereby hindering metaphorical abstraction. Third, multisensory integration is impaired in schizophrenia (de Gelder et al., 2003; Tseng et al., 2015; Williams et al., 2010), as supported, for instance, by evidence from the rubber-hand illusion paradigm (Prikken et al., 2019; Zopf et al., 2021). Patients with schizophrenia are more susceptible to the illusion than healthy controls, suggesting an atypical weighting of visual over proprioceptive information. This altered balance might be particularly relevant for figurative language, where visual and verbal cues must be flexibly integrated to derive abstract meanings (Tonna et al., 2023). Importantly, recent work showed an association between greater susceptibility to bodily illusions and poorer metaphor comprehension in schizophrenia (Battaglini et al., 2026) and introduced the idea that metaphor impairment might arise from overweighing perceptual activations relative to contextual constraints during figurative meaning constructions.

To test the hypothesis that the overactivation of visual representations evoked by metaphors contributes to difficulties in figurative language comprehension in schizophrenia, this study pursued two complementary aims: i) to establish altered sensory-motor activation during metaphor comprehension in patients compared to control, in particular overactivation of visual properties; ii) to test whether such overactivation is predictive of accuracy in metaphor comprehension, after controlling for well-known predictors such as verbal and cognitive abilities. For aim i), we developed a novel metaphor priming experiment where metaphors (e.g., *Wisdom is a flashlight*) were used as primes for words related to the metaphor vehicle (*flashlight*) based on semantic (*lamp*), visual (*microphone*), or action (*remote*) properties in a lexical decision task (Figure 1). Target words were presented at different inter-stimulus intervals (ISIs), to test the temporal dynamics of priming effects. Previous studies have shown that, in healthy participants, words can prime other words related through visual or motor properties (e.g., *soldering iron* or *housekey* can prime *screwdriver*; Lam et al., 2015). More generally, priming paradigms have proved particularly suitable for probing automatic bottom-up activation while dissociating it from top-down predictive influences (Tumanyants & He, 2025). Likewise, metaphors can prime not only semantically related words (Rubio Fernández, 2007) but also words that are related based on sensory-motor properties (Al-Azary & Katz, 2021), which has been taken as evidence for the activation of those sensory-motor features. Hence, in line with our hypothesis of visual overactivation in schizophrenia, we expected enhanced and/or longer-lasting metaphor priming for visually related words in patients compared to controls. Conversely, given the evidence of reduced semantic priming in schizophrenia (Minzenberg et al., 2002; Pomarol-Clotet et al., 2008) and the hypothesis of underweighted contextual priors, we expected weaker and/or more transient priming effects for semantically related words in patients compared to healthy controls. For action priming, the evidence in the literature is mixed, reporting both facilitation and interference effects between figurative language and motor processing (Garello et al., 2024; Wilson & Gibbs, 2007); therefore, no specific predictions were formulated, but action-related targets were nevertheless included to explore potential abnormalities in metaphor processing extending beyond the visual domain. For aim ii), we collected independent measures of metaphor comprehension accuracy and general verbal and cognitive skills, and we tested whether metaphor priming effects observed in the priming experiment were predictive of accuracy in a separate multiple-choice metaphor comprehension task, controlling for other factors. In addition, we investigated whether priming effects varied as a function of metaphor familiarity and metaphor concreteness, given the evidence of different embodiment processes depending on item types even in neurotypical populations (Cacciari et al., 2011; Romero Lauro et al., 2013).

**Figure 1.**
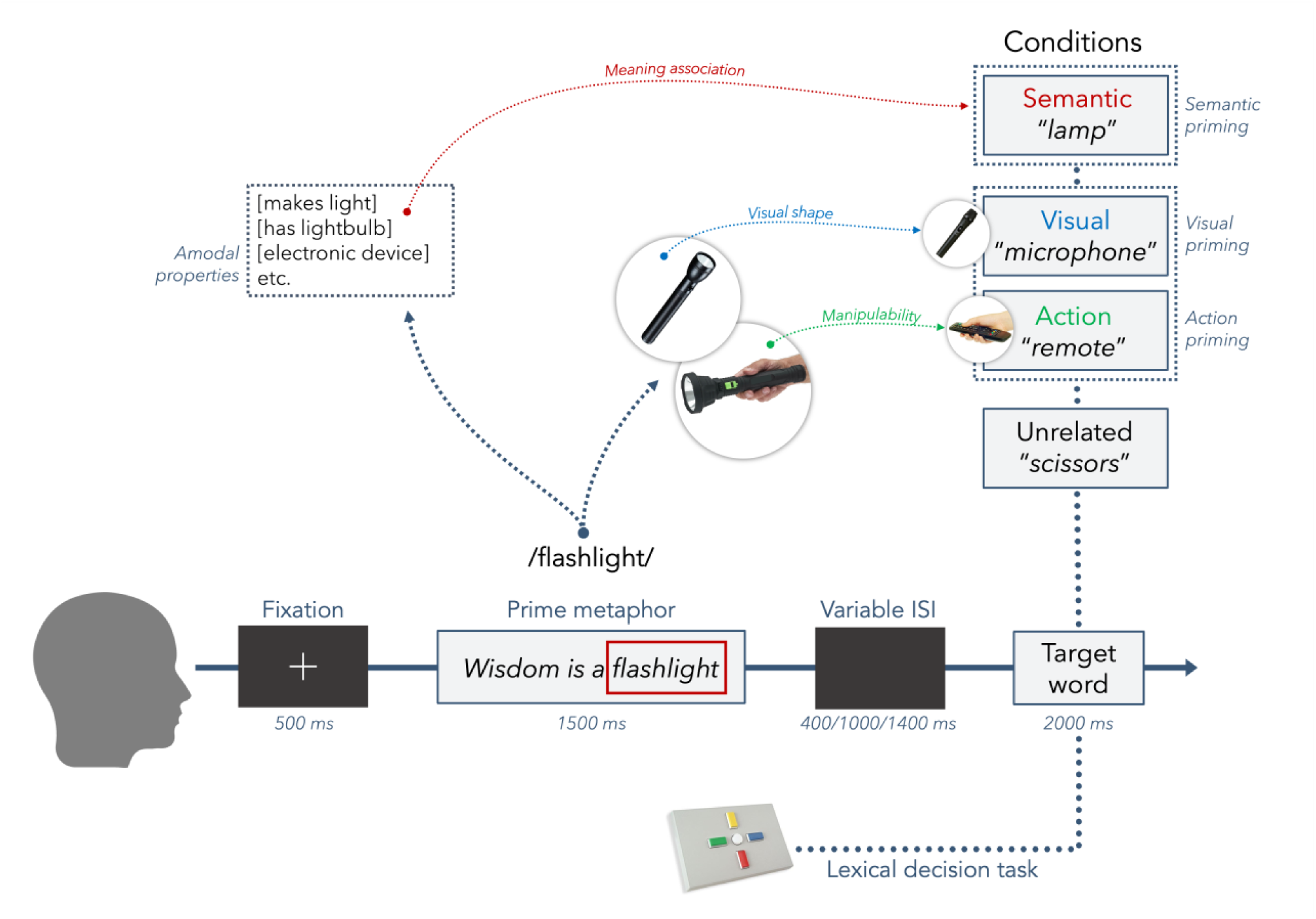
The structure of the metaphor priming experiment. Participants read a metaphorical prime (e.g., “Wisdom is a flashlight”, with “flashlight” as the metaphor vehicle) followed, after a variable inter-stimulus interval (ISI), by a target words. Target words could be unrelated with respect to the metaphor vehicle (i.e., “scissors”) or associated with it by either amodal semantic properties (i.e., meaning association, as with “lamp”) or modal properties, such as visual (i.e., visual shape, as with “microphone”) or action (i.e., manipulability, as with “remote”) characteristics. Participants were asked to perform a lexical decision task, indicating whether the target was an existing Italian word by pressing one of two buttons.

## 2. Results

### 2.1. Demographic, clinical, and cognitive characteristics

The clinical sample included 66 patients with schizophrenia, who were all stabilized and treated with antipsychotic therapy for at least 3 months. The control group included 70 participants, balanced for age and education with the schizophrenia group. Descriptive statistics for demographic, clinical, and cognitive measures as well as between-group comparisons are reported in Table 1.

**Table 1.** Descriptive statistics (mean and standard deviations) of patients and controls with group comparisons across demographic, clinical, and cognitive measures.

| Measures | Patients<br>Mean (SD) | Controls<br>Mean (SD) | Test Statistics | p-value |
| --- | --- | --- | --- | --- |
| Age | 40.03 (13.01) | 42.13 (15.42) | $t(134) = 0.86$ | $p = .394$ |
| Education (years) | 11.62 (2.69) | 11.99 (2.61) | $t(134) = 0.80$ | $p = .424$ |
| Gender (F/M) | 17/49 | 50/20 | $\chi^2(1) = 28.35$ | $p < .001$ |
| Illness onset (years) | 23.08 (6.20) | - | - | - |
| Illness duration (years) | 16.95 (10.53) | - | - | - |
| Mean chlorpromazine-equivalent dose (mg/d) | 493.55 (227.80) | - | - | - |
| PANSS Positive | 16.98 (4.75) | - | - | - |
| PANSS Negative | 21.61 (6.38) | - | - | - |
| PANSS General | 40.37 (9.20) | - | - | - |
| PANSS Disorganization | 21.17 (6.44) | - | - | - |
| WAIS-R Vocabulary | 44.86 (11.6) | 52.22 (9.72) | $t(131) = 3.97$ | $p < .001$ |
| APACS Brief Total | 0.61 (0.11) | 0.87 (0.08) | $t(59.61) = 12.81^b$ | $p < .001$ |
| BACS<sup>a</sup> | 1.70 (0.92) | 2.63 (0.71) | $t(112.07) = 6.46$ | $p < .001$ |
| ToM-PST<sup>c</sup> | 48.63 (9.33) | - | - | - |
| QMI Total | 53.48 (8.21) | 61.00 (6.97) | $t(132) = 5.73$ | $p < .001$ |
| MIT Total | 14.70 (8.21) | 15.01 (6.97) | $t(132) = 0.94$ | $p = .351$ |
| PWB Total | 159.29 (22.20) | 193.87 (16.20) | $t(124.43) = 10.34$ | $p < .001$ |
| Metaphor<br>Comprehension Total | 16.30 (2.41) | 17.34 (3.42) | $t(115.97) = 2.04$ | $p = .044$ |

Notes. PANSS = Positive and Negative Syndrome Scale, BACS = Brief Assessment of Cognition in Schizophrenia, ToM-PST = Theory of Mind Picture Sequencing Task, QMI = Questionnaire upon Mental Images, MIT = Mental Imagery Test, PWB = Psychological Well-Being Scales. All p-values were adjusted for False Discovery Rate (FDR).
<sup>a</sup> Mean of the equivalent scores from each subtask of the BACS
<sup>b</sup> The comparison refers to 108 participants only, as the test could not be administered to other participants
<sup>c</sup> This measure was available for a subset of patients only ( $n = 52$ )

The *t*-tests comparing the two groups showed that participants with schizophrenia scored significantly worse than control participants in lexical-semantic skills (as assessed with the WAIS-R Vocabulary subtask), pragmatic skills (as assessed with the APACS Brief test), and in neurocognition (as assessed via the BACS). Participants with schizophrenia reported lower vividness of mental images, as evaluated using the QMI, but did not score lower than controls in tasks assessing manipulability and maintenance of mental images (as evaluated with MIT tasks). Participants with schizophrenia reported reduced psychosocial well-being compared to control subjects, as indicated by the PWB scales, and showed worse performance in the multiple-choice metaphor comprehension task.

### 2.2. Results of the metaphor priming experiment

Raw latencies for the responses to the lexical decision task included in the metaphor priming experiment across the different conditions (unrelated, semantic, visual, action) are displayed in Table 2. Semantic priming effects, i.e., facilitation on semantically related words compared to the unrelated baseline, along with Visual and Action priming effects, i.e., facilitation on visually and action-related words compared to the semantic baseline, exerted by the metaphors in the lexical decision task are also shown in Table 2 and depicted in Figure 2.

**Figure 2.**
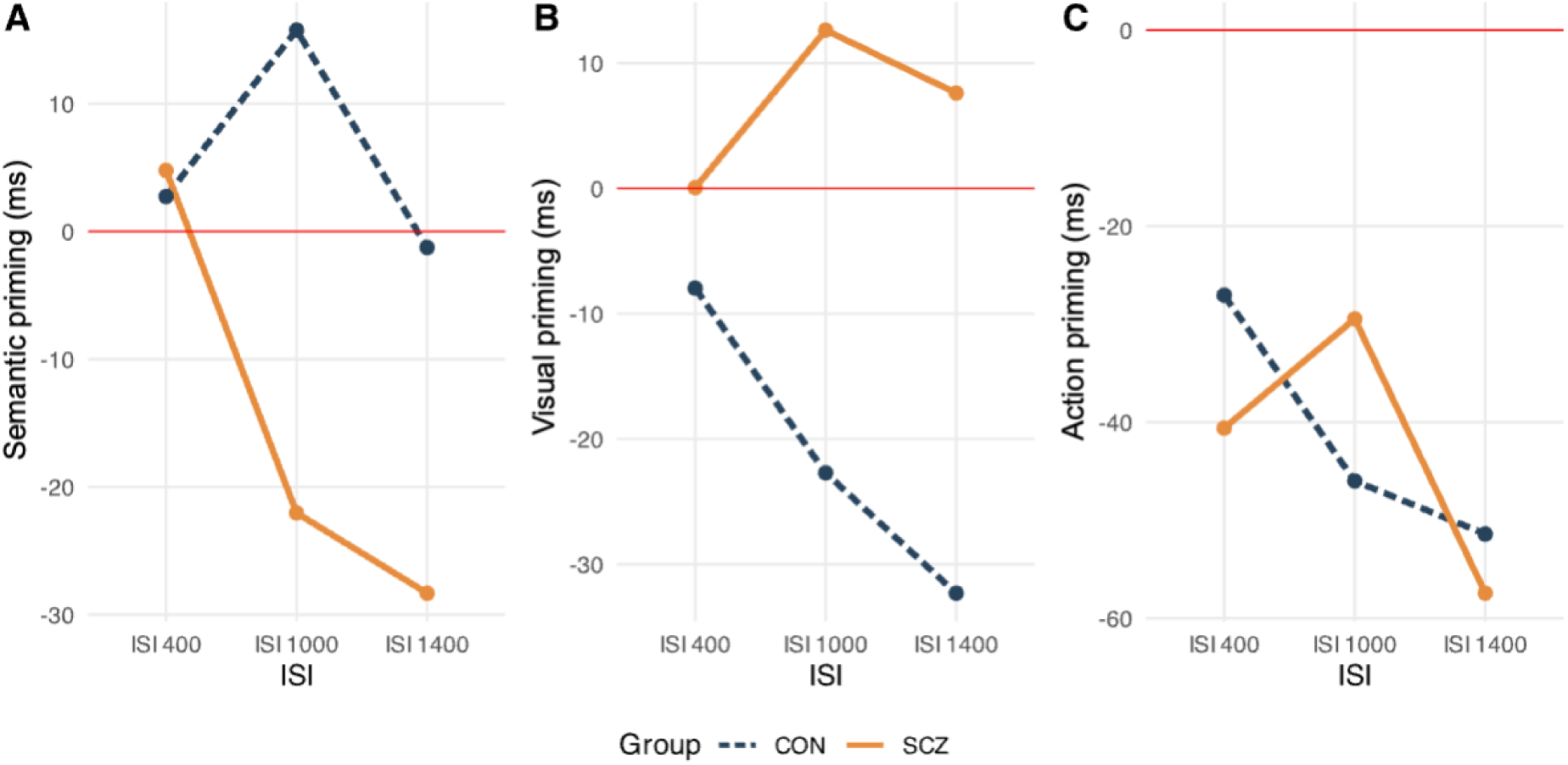
Semantic, Visual and Action priming effects across groups and inter-stimulus intervals (ISI). Mean semantic (A), visual (B), and action (C) priming effects (in milliseconds) for schizophrenia (SCZ) and control (CON) groups, across 400, 1000, and 1400 millisecond ISIs. Positive and negative values indicate facilitatory and inhibitory effects of the condition relative to the baseline, respectively. The red line indicates the absence of priming effects.

**Table 2.** Latencies in the metaphor-primed lexical decision task across conditions and inter-stimulus intervals (ISI) and metaphor priming effects in patients and controls.

| Condition | ISI 400 ms |  | ISI 1000 ms |  | ISI 1400 ms |  |
| --- | --- | --- | --- | --- | --- | --- |
|  | Controls | Patients | Controls | Patients | Controls | Patients |
| Unrelated | 728.86<br>(190.71) | 859.14<br>(306.96) | 781.08<br>(241.61) | 811.84<br>(260.79) | 735.85<br>(186.48) | 811.65<br>(208.42) |
| Semantic | 726.12<br>(228.88) | 854.36<br>(297.30) | 765.32<br>(214.89) | 833.87<br>(264.40) | 737.10<br>(201.91) | 839.97<br>(238.43) |
| Visual | 734.09<br>(255.55) | 854.30<br>(300.09) | 788.03<br>(230.42) | 821.25<br>(266.32) | 769.41<br>(227.77) | 832.36<br>(234.93) |
| Action | 753.17<br>(213.56) | 894.95<br>(334.48) | 811.31<br>(223.64) | 863.33<br>(266.89) | 788.50<br>(231.10) | 897.40<br>(251.44) |
| Semantic Priming | 2.73 | 4.77 | 15.76 | -22.03 | -1.25 | -85.75 |
| Visual priming | -7.96 | 0.06 | -22.71 | 12.63 | -32.31 | 7.60 |
| Action priming | -27.04 | -40.58 | -45.99 | -29.45 | -51.39 | -57.43 |

*Notes. Mean and standard deviations (in brackets). All values are in milliseconds.*

The Linear Mixed-Effects model testing semantic, visual, and action priming effects (Table 3) showed a main effect of Group at 400 ms and – marginally – at 1400 ms, indicating that in these time windows, patients with schizophrenia were globally slower than controls in the lexical decision task. Across ISIs, the model also showed a main negative effect of Condition for action priming effects, with slower responses in the action compared to the semantic condition in both groups, indicating that for all participants, metaphors hindered response to action-related target words. Two significant interactions involving Group emerged. Specifically, we observed a Group × Condition interaction for the semantic priming at 1000 ms: in this time window, healthy controls showed faster responses to semantic target words (relative to the unrelated) compared to patients, indicating that in healthy controls, metaphors facilitated responses to semantically related target words relative to baseline, reflecting greater activation of semantic properties of the metaphor vehicle. The inspection of the data evidenced that, while controls showed a semantic priming pattern exerted by the metaphors, patients actually exhibited reverse priming (i.e., negative priming values), with metaphors inhibiting the response to semantically related words in patients. The model also detected a significant Group × Condition interaction for the visual priming at 1000 and 1400 ms: in these time windows, the schizophrenia group showed faster responses to the visual target words (relative to the semantic) compared to the control group, indicating that in patients, metaphors facilitated responses to visually related target words relative to baseline, reflecting greater activation of visual properties of the metaphor vehicle. The inspection of the data evidenced that, while patients showed a visual priming pattern exerted by the metaphors, controls actually exhibited a reverse priming (i.e., negative priming values), with metaphors inhibiting the response to visually related words in controls.

**Table 3.**
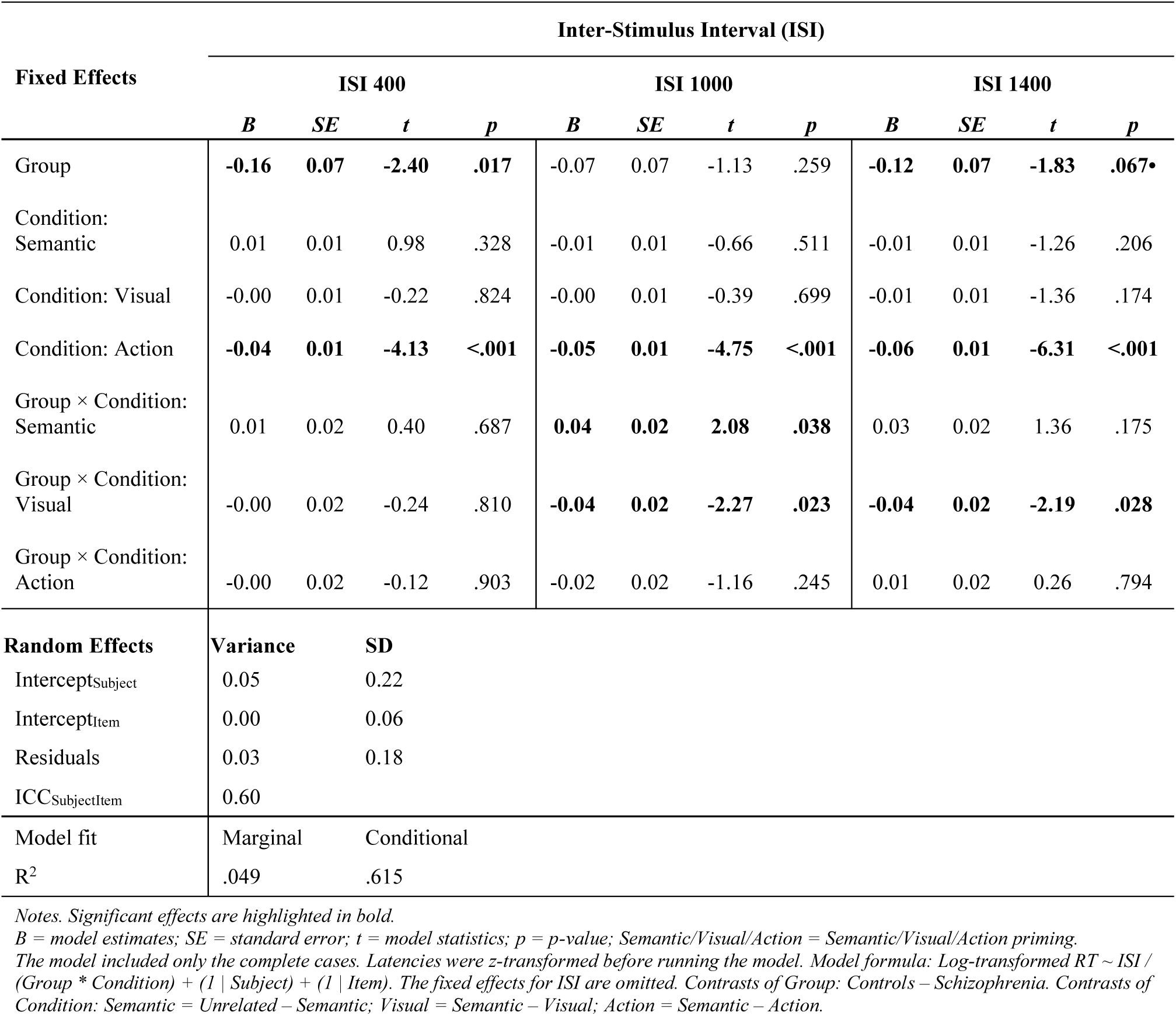
Output of the Linear Mixed-Effects Model testing the presence of semantic, visual and action priming effects across inter-stimulus intervals (ISI) in the schizophrenia and control groups.

### 2.3. Relation with metaphor comprehension, individual and item differences

When correlating the significant priming effects (averaged by subjects) with the accuracy score of the metaphor comprehension task (Figure 3A), we found that, in patients, visual priming was negatively associated with metaphor comprehension scores at 1000 ms (but not at 1400 ms), indicating that patients with higher activations of visual properties of the metaphor vehicle at 1000 ms exhibited poorer metaphor comprehension. Additionally, metaphor comprehension in patients was negatively associated with age and with several PANSS dimensions, and positively with education, WAIS-R Vocabulary total score, the BACS Average Equivalent Score, and ToM-PST total score, indicating that metaphor comprehension was poorer in patients with older age and greater psychopathological severity, and better in those with higher education and stronger lexical-semantic, cognitive, and sociocognitive skills.

**Figure 3.**
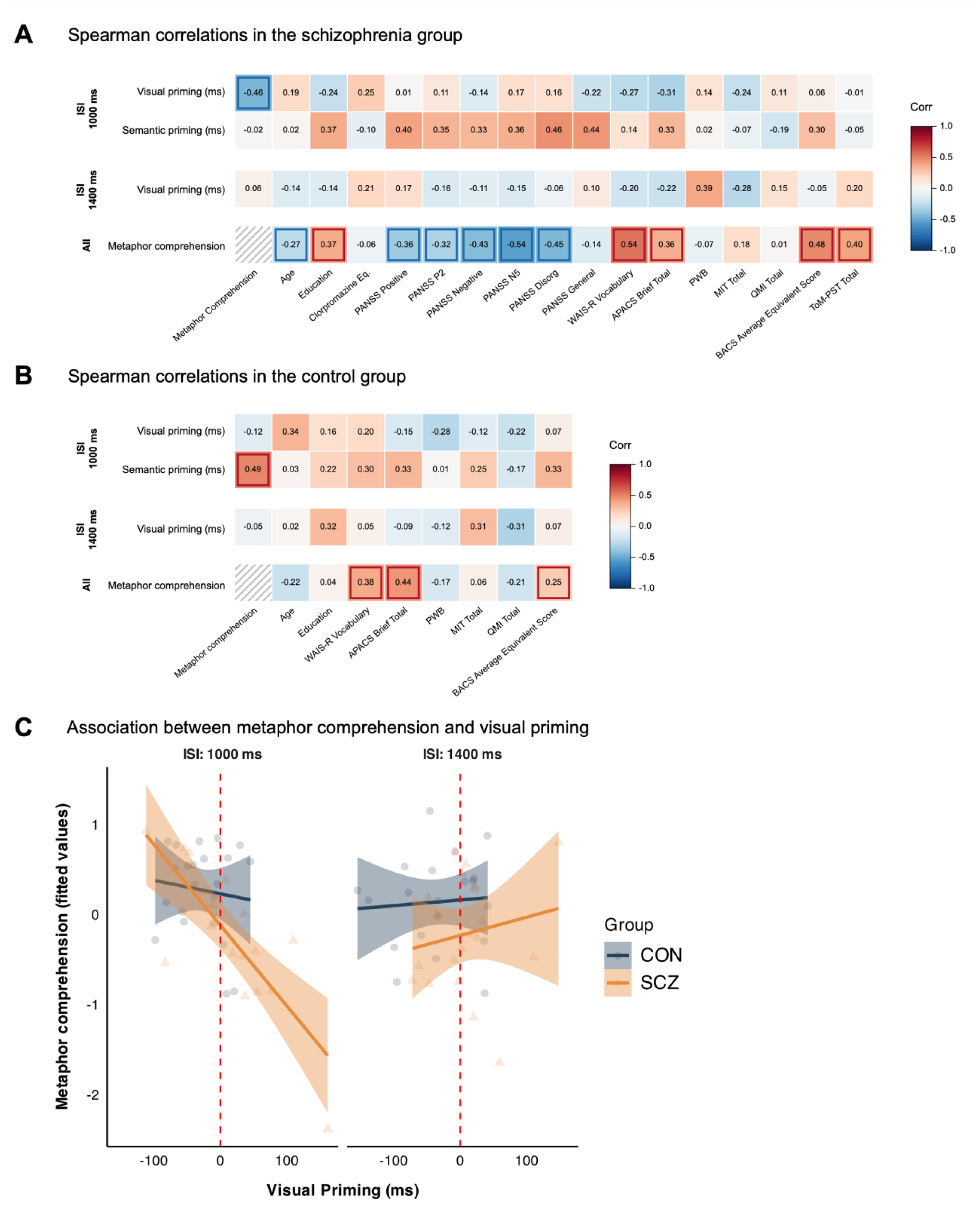
Associations between metaphor priming effects and assessment measures. **(A-B)** Spearman’s correlations between metaphor-induced semantic and visual priming effects (mean reaction time differences, in ms) and demographic, clinical, and cognitive variables in schizophrenia and control groups, separately for 1000 and 1400 ms inter-stimulus interval (ISI) subgroups (metaphor comprehension correlations was based on all participants). Color indicates correlation strength and direction, with significant cells (p < .05) highlighted. (C) Scatterplots illustrating the association between visual priming and metaphor comprehension in schizophrenia (SCZ) and control (CON) participants as resulting from the multiple regression model, separately for 1000 and 1400 ms ISI subgroups. Note: Metaphor Comprehension = total score of the multiple-choice metaphor comprehension task; PANSS = Positive and Negative Syndrome Scale; BACS = Brief Assessment of Cognition in Schizophrenia; PWB = Psychological Well-Being Scales; MIT = Mental Imagery Test; QMI = Questionnaire upon Mental Images; ToM-PST = Theory of Mind Picture Sequencing Test.

Conversely, in healthy controls (Figure 3B), semantic priming effects were positively associated with metaphor comprehension at 1000 ms, indicating that individuals with higher activations of semantic properties of the metaphor vehicle at 1000 ms achieved better metaphor comprehension. Additionally, metaphor comprehension in controls was positively associated with WAIS-R Vocabulary and with the BACS Average Equivalent Score, indicating that metaphor comprehension was higher in individuals with stronger lexical-semantic and neurocognitive skills.

Notably, Priming effects did not correlate with any other measure in either group, suggesting that susceptibility to metaphor-induced priming reflected a relatively specific processing tendency rather than a consequence of cognitive impairment or symptom severity.

When testing in a more stringent fashion whether priming effects predicted metaphor comprehension independently of lexical-semantic and neurocognitive skills via multiple regressions (see Table 4 and Figure 3C), we found that in the schizophrenia group, visual priming at 1000 ms (but not at 1400 ms) remained a significant predictor of metaphor comprehension, while lexical-semantic and neurocognitive skills were only marginally associated with performance. No variable significantly predicted metaphor comprehension in healthy controls.

**Table 4.** Output of the multiple regression testing the association between the Metaphor Comprehension Task and the semantic and visual priming effects across groups and relevant inter-stimulus intervals (ISI), controlling for lexical-semantic and neurocognitive abilities.

| Predictors | ISI | Group | *B* | *SE* | *t* | *p* |
| --- | --- | --- | --- | --- | --- | --- |
| WAIS-R Vocabulary (Total score) |  |  | 0.30 | 0.16 | 1.88 | .064• |
| BACS Average Equivalent Score |  |  | 0.28 | 0.15 | 1.91 | .061• |
| Mean Semantic Priming | 1000 ms | SCZ | -0.26 | 0.28 | -0.92 | .362 |
| Mean Visual Priming |  |  | -0.53 | 0.23 | -2.33 | **.023** |
| Mean Semantic Priming |  | CON | 0.15 | 0.20 | 0.75 | .454 |
| Mean Visual Priming |  |  | -0.22 | 0.31 | -0.71 | .482 |
| Mean Semantic Priming | 1400 ms | SCZ | -0.25 | 0.22 | -1.12 | .266 |
| Mean Visual Priming |  |  | 0.12 | 0.24 | 0.52 | .603 |
| Mean Semantic Priming |  | CON | 0.18 | 0.19 | 0.90 | .372 |
| Mean Visual Priming |  |  | 0.05 | 0.20 | 0.25 | .803 |
| Model fit | R<sup>2</sup> | R<sup>2</sup> adjusted |  |  |  |  |
|  | .402 | .282 |  |  |  |  |

Notes. Significant effects are highlighted in bold
*B* = model estimates; *SE* = standard error; *t* = model statistics; *p* = *p*-value; ISI = inter-stimulus interval; CON = controls, SCZ = schizophrenia.
The model included priming scores aggregated by participants using only the complete cases. Two subjects from the control group were removed from the model as they showed highly deviant values in the dependent variable. Model formula: Metaphor Comprehension (Total) ~ WAIS-R Vocabulary (Total) + BACS Average Equivalent Score + ISI / Group / Mean Semantic Priming + Mean Visual Priming. All continuous variables (including the dependent variable) were z-transformed before entering the model. The main effects for ISI and Group are omitted.

When correlating the significant priming effects (averaged by item) with item characteristics, we found that visual priming was positively correlated with the concreteness of the metaphor topic (*r*(18) = .57, *p* = .009), indicating that more concrete metaphors exerted stronger visual priming effects (Figure 4A), but not with metaphor familiarity (*r*(18) = -.13, *p* = .599). Conversely, semantic priming showed a marginal positive correlation with metaphor familiarity (*r*(18) = .43, *p* = .063), indicating that more familiar metaphors exerted stronger semantic activations (Figure 4B), but not with the concreteness of the topic (*r*(18) = .18, *p* = .456).

**Figure 4.**
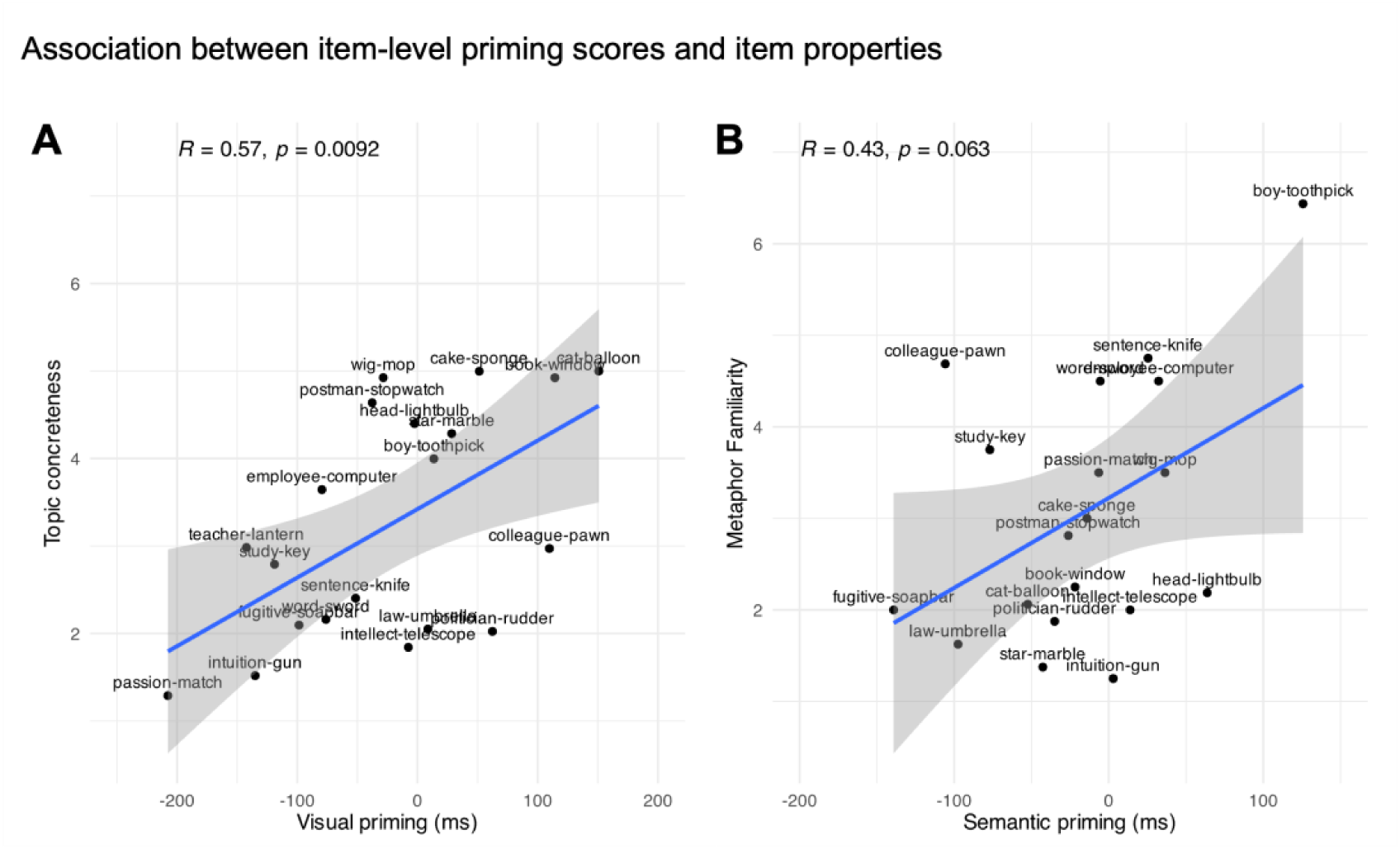
Associations between item-level metaphor priming effects and metaphor properties. Pearson’s correlations between visual priming effects (aggregated by item) and concreteness of the metaphorical prime topic (A), and between semantic priming effects (aggregated by item) and metaphor familiarity (B). Each point represents one metaphor, labelled by its topic–vehicle pair.

## 3. Discussion

In this study, we tested the novel hypothesis that figurative language impairment in schizophrenia may partly arise from altered perceptual processing during metaphor comprehension, and specifically from the overactivation of visual representations evoked by metaphorical expressions. To this end, we developed a metaphor priming paradigm where metaphors were used as primes for words associated with the metaphor vehicle based on semantic, visual, or motor characteristics. We expected to detect stronger or longer-lasting priming effects in schizophrenia for visually-related target words, consistent with the idea that patients are more prone to activate and maintain the visual representations evoked by metaphors, and to observe a genuine link between such activations and patients’ difficulties in understanding metaphors. The results largely aligned with our hypotheses, by showing that, compared to controls, patients with schizophrenia exhibited sustained visual priming effects, i.e., facilitation for visually related target words, emerging 1000 ms after metaphor presentation and persisting up to 1400 ms. Importantly, stronger metaphor-induced visual priming predicted poorer scores in the metaphor comprehension task, even after controlling for neurocognitive and vocabulary skills, indicating that patients showing higher activation of visual properties associated with the metaphor vehicle were also those with poorer metaphor comprehension. Conversely, healthy controls showed semantic priming effects at 1000 ms, which positively correlated with metaphor comprehension ability. Taken together, these findings support the view that metaphor comprehension in schizophrenia is characterized by an altered activation of visual information and an imbalance between perceptual and semantic processing streams.

As a first consideration, the present findings allow us to extend classic accounts of concretism in schizophrenia by clearly highlighting the role of perceptual processing. While traditional cognitive or psychopathological accounts defined concretism as the inability to move beyond immediately available stimuli (Harrow, 1974), we showed that, in the case of metaphor understanding, concretism is specifically associated with perceptual processing disruption. After being presented with a metaphor such as *Wisdom is a flashlight*, individuals with schizophrenia appear to activate visual representations linked to the metaphor vehicle in its literal sense (e.g., the image of a flashlight or some of its visual properties) and to maintain such representations over time. For healthy controls, conversely, there is no trace of sustained visual activations at these latencies, with the metaphor actually becoming an obstacle for recognizing visually related words (as indicated by the negative priming values). Far from excluding the involvement of perceptual simulation in neurotypical individuals, it is possible that it contributes to the initial stages of figurative interpretation, as attested, for instance by N400 effects modulated by metaphor concreteness (Canal et al., 2022), becoming subsequently integrated into more abstract conceptual representations and no longer exerting measurable effects after the end of the metaphor. Overall, the present findings are consistent with previous evidence indicating that patients with schizophrenia tend to use more concrete words compared to controls when explaining figurative expressions (Bambini et al., 2025) and that concrete features of words are abnormally activated in schizophrenia at relatively late latencies (Spitzer, 1993, 1997). We refined these accounts by showing that the critical alteration specifically concerns visual-perceptual representations, which are maintained over time and are directly related to metaphor comprehension difficulties.

To better understand the pattern of results, it is useful to discuss in detail also the semantic priming effects reported in healthy participants, namely the facilitation of semantically related words. Following the presentation of a metaphor such as *Wisdom is a flashlight*, healthy controls appear to maintain activation of a network of semantically associated concepts (e.g., *lamp*), likely containing more abstract semantic properties coherent with the figurative meaning of the expression, such as guidance or clarity. At these latencies, while visual representations no longer facilitate processing and may even interfere with it, semantically associated terms remain active to support figurative interpretation. Patients, conversely, showed negative semantic priming values. This can be related to the hypopriming effects frequently reported in schizophrenia at relatively long latencies (Kreher et al., 2009; Kuperberg, 2010; Sass et al., 2014), but it may also point to more specific difficulties in maintaining and integrating abstract semantic representations after metaphor presentation. In this sense, our results extend the idea of concretism not only in the direction of a link with perceptual processing alteration, but also in terms of integration between perceptual and semantic information. Item-level analyses further support the distinction between visual and semantic routes in metaphor processing. Visual priming was stronger for metaphors with more concrete topics (such as, for instance, *That cat is a balloon*), suggesting that metaphors grounded in more imageable concepts may engage perceptual representations more strongly. By contrast, semantic priming tended to increase for more familiar metaphors (e.g., *That boy is a toothpick*), consistent with the idea that repeated exposure facilitates the emergence of more abstract and semantically integrated representations. This pattern suggests that visual and semantic activations may reflect partially distinct pathways in metaphor comprehension, one more closely tied to perceptual simulation and the other to semantic knowledge, in line with the dual coding account (Canal et al., 2022; Paivio, 1979; Paivio & Walsh, 1993) and, more broadly, with a multimodal account of metaphor (Battaglini et al., 2025; Frau, Tomasello, et al., 2026). In healthy individuals, these routes may be flexibly coordinated during figurative interpretation, whereas in schizophrenia an altered balance between perceptual and semantic processing may contribute to concrete interpretations.

The correlational analysis further complements this picture. Beyond demonstrating the link between visual priming and metaphor comprehension performance, correlations also highlighted the broader constellation of cognitive and linguistic abilities involved in figurative interpretation. Consistent with previous literature (Bambini et al., 2016; Frau et al., 2024; Mossaheb et al., 2014; Parola et al., 2018), metaphor comprehension in patients was associated with lexical-semantic abilities, cognitive performance, and Theory of Mind measures, suggesting that successful metaphor processing likely depends on multiple interacting mechanisms. Crucially, however, the effect of visual priming on metaphor comprehension remained significant also when controlling for neurocognitive and verbal skills, as shown in the regression model. Moreover, priming effects themselves were not associated with any neuropsychological tests, suggesting that overactivation of visual information does not simply reflect generalized cognitive impairment, but may instead index an altered lower-level regulation of perceptual activity. Importantly, priming effects were also unrelated to explicit measures of imagery, such as the QMI and the MIT. In the QMI questionnaire, patients reported lower scores relative to controls, possibly because of the metacognitive demands associated with the tasks (Pearson et al., 2011), while in the MIT tasks they performed as the control subjects. The dissociation between imagery measures and visual priming is consistent with the different processes captured by the two paradigms. Whereas imagery questionnaires assess the conscious generation of mental images (Frau, Bischetti, et al., 2025; Muraki & Pexman, 2021), priming paradigms tap into the automatic activation of conceptual representations during language comprehension. Although related, these forms of visual processing are at least partly distinct (Muraki et al., 2023). The observed visual priming effects, therefore, are more likely to reflect abnormalities in the involuntary activation and maintenance of visual representations evoked by metaphorical expressions than differences in voluntary imagery.

More broadly, these findings can be situated within several broader physiological abnormalities described in schizophrenia. First, they resonate with evidence of impaired sensory gating, namely a reduced ability to filter or suppress irrelevant sensory information (Javitt & Freedman, 2015; Silverstein & Keane, 2011). Likewise, growing evidence points to abnormalities in the integration of sensory signals across modalities, as illustrated by altered responses in paradigms such as the rubber-hand illusion (Prikken et al., 2019; Zopf et al., 2021). Converging evidence also comes from mismatch negativity (MMN), one of the most robust neurophysiological markers of schizophrenia (Kim et al., 2020; Lee et al., 2014), which has been interpreted as reflecting attenuated sensory prediction and has recently been linked to pragmatics (Agostoni et al., 2026). Against this background, the present findings provide a possible neurophysiological mechanism linking these broader abnormalities in sensory processing to figurative language impairment. The predictive-processing account offers a unifying framework for interpreting this mechanism. Within this view, schizophrenia has been proposed to involve altered precision-weighting mechanisms, whereby bottom-up sensory signals receive excessive weight relative to higher-order contextual constraints (Corlett et al., 2019; Sterzer et al., 2018; Weilnhammer et al., 2020). Notably, this framework has already been extended to language, where linguistic disorganization has been proposed to arise from a failure of top-down contextual guidance (Palaniyappan, 2022). Applied to metaphor comprehension, the present findings suggest that visual representations associated with the metaphor vehicles are differently treated in patients and healthy individuals. In healthy controls, perceptual information, when activated, is downregulated, as contextual and semantic constraints guide interpretation toward the figurative meaning, consistent with recent evidence that predictive processes contribute to metaphor processing (Lago et al., 2024). This pattern is reflected in the reverse visual priming together with semantic priming observed in controls. In schizophrenia, by contrast, perceptual representations may receive excessive weight relative to contextual priors, resulting in prolonged visual activation together with interference for semantic processing. Under this view, concretism may emerge not simply from a failure of abstraction but from a difficulty in appropriately regulating perceptual information during meaning construction. Similar mechanisms have been proposed to explain abnormalities in domains other than language, including face perception and perceptual organization, where excessive influence of sensory information and reduced top-down modulation have been linked to altered interpretation of stimuli (Butler et al., 2008; Silverstein & Keane, 2011).

Interestingly, neither patients nor controls showed priming effects for action-related target words. This finding suggests that motor simulations may play a less stable or less central role in figurative processing, becoming relevant only in specific conditions, for instance when bodily actions are directly coherent with the intended figurative interpretation itself (Al-Azary & Katz, 2021), or at earlier stages of processing (Tomasello, 2023). Previous studies have indeed reported both facilitation and interference effects between figurative language and motor processing, suggesting a more context-dependent contribution of motor simulation (Cuccio et al., 2014; Garello et al., 2024).

Several limitations should be acknowledged. First, although the present results identify a relationship between visual priming and metaphor comprehension difficulties, the study design does not allow direct conclusions regarding causality. Future studies employing neurophysiological or neuromodulatory approaches may help clarify whether abnormal perceptual activations play a mechanistic role in concretistic interpretation. Second, the present findings concern metaphors specifically, and future work should examine whether similar perceptual-processing abnormalities extend to other forms of figurative language, such as idioms or proverbs, and to expressive language skills, which are also impaired in schizophrenia (Meister et al., 2026). For instance, whether abnormalities in bottom-up perceptual processing contribute to incoherence and tangentiality remains an open question.

Overall, the present study provides evidence that figurative language impairment in schizophrenia is not solely related to deficits in executive or social-cognitive processes but may also involve altered dynamics within perceptual systems. By linking metaphor comprehension difficulties with overactivation of visual representations, these findings support a novel account of concretism grounded in low-level visual mechanisms and in their abnormal integration with semantic processing during language comprehension. More broadly, these findings suggest that concretism may reflect a failure to appropriately regulate perceptually grounded representations during language comprehension, providing a novel mechanistic framework linking embodied cognition, predictive processing, and pragmatic impairment in schizophrenia. We speculate that this novel mechanistic account could lead to innovative treatments for language disorders in schizophrenia aimed at restoring the balance between bottom-up perceptual signals and higher-order interpretative processes. This would extend the main strategy of current pragmatic rehabilitation programs (Bambini, Agostoni, et al., 2022), mainly based on improving attention to context, with a perceptual-flexibility module targeting the dynamic integration of visually grounded and contextual information during figurative language comprehension.

## 3. Methods

### 3.1. Participants

Before running the study, we performed an *a priori* power analysis using G*Power software (Faul et al., 2009) to estimate the optimal sample size for this study: in line with moderate-to-large effect sizes reported by Lam et al. (2015) in a similar priming experiment (0.20 ≤ *f^2^*s ≤ 0.33, determined following Lakens, 2013), the power analysis indicated the required sample size to achieve 80% power for detecting a medium effect reflecting a significant difference between target conditions (*f*^2^ ≥ 0.15, Cohen, 1988), at a significance criterion of *α* = .05, was *N* = 114, accounting for the potential exclusion of participants during data cleaning (∼15% of subjects). Accordingly, we recruited a total sample of 143 participants belonging to either the clinical or the control groups.

The clinical group included 70 individuals with a diagnosis of schizophrenia based on DSM-5 criteria (American Psychiatric Association, 2013), recruited from the Department of Clinical Neurosciences, IRCCS San Raffaele Scientific Institute, Milan, Italy. Inclusion criteria comprised being: a) a native speaker of Italian; b) within the age range of 18–65 years; c) clinically stabilized and treated with a stable dose of the same antipsychotic therapy for at least 6 months. Exclusion criteria were: a) severe traumatic brain injury or neurological disorders; b) intellectual disability; c) alcohol or substance abuse in the preceding 6 months; d) severe psychotic exacerbation in the preceding 3 months. Four participants were excluded as they met one of the exclusion criteria.

Additionally, 73 Italian-speaking healthy controls, balanced for age and education with the sample of patients, were recruited from different regions of Italy. Inclusion criteria for the control group were being: a) a native speaker of Italian; b) within the age range of 18–65 years. Participants were excluded if they had a diagnosis of psychiatric or neurological disorders. One control subject was excluded from the sample, while two other control subjects dropped out of the study and were not included in the sample.

All participants provided informed consent. The study was approved by the local ethical committees (RS: 318/06 “Studio sulla qualità della vita dei pazienti psicotici in funzione delle caratteristiche cliniche e biologiche della malattia”; Department of Brain and Behavioral Sciences of the University of Pavia: 122/23 “Meta-Imagery: Il ruolo delle immagini mentali visuo-motorie nella comprensione delle metafore e nel disturbo pragmatico (PROMENADE, WP4 “Decay”, WP4.2)”), following the principles of the Declaration of Helsinki.

### 3.2. Assessment

All participants completed a comprehensive assessment battery evaluating vocabulary, pragmatic abilities, neurocognitive functioning, psychosocial well-being, and mental imagery. Participants in the schizophrenia group underwent additional clinical assessment of psychopathology and theory of mind skills conducted by a trained psychiatrist. To minimize fatigue, patients were assessed across three sessions of approximately one hour each, whereas controls completed the protocol in a single session lasting approximately 90 minutes, including a short break.

Psychopathology was assessed using the Positive and Negative Syndrome Scale (PANSS; Kay et al., 1987), which measures positive, negative, and general psychopathological symptoms. In line with previous studies (Bambini, Frau, et al., 2022; Lucarini et al., 2022), a disorganization composite score was also computed.

ToM abilities were evaluated using the ToM Picture Sequencing Task (ToM-PST; Brüne, 2003). In this task, participants are required to arrange picture cards into a coherent sequence and answer questions concerning the characters’ mental states. The task yields a total score ranging from 0 to 59. Vocabulary knowledge was assessed with the Italian version of the WAIS–R Vocabulary subtest (Orsini & Laicardi, 1997), in which participants provide definitions for 35 words of increasing difficulty. Total scores range from 0 to 70.

Pragmatic abilities were measured using the brief version of the Assessment of Pragmatic Abilities and Cognitive Substrates test (APACS Brief; Bischetti et al., 2025; Bischetti, Pompei, et al., 2024), which evaluates expressive and receptive pragmatic skills through five subtests (Interview, Narratives, Figurative Language 1 and 2, and Humor), yielding a standardized total score ranging from 0 to 1.

Neurocognitive functioning was assessed using the Italian version of the Brief Assessment of Cognition in Schizophrenia (BACS; Anselmetti et al., 2008). The battery comprises seven tasks assessing six cognitive domains: verbal memory, working memory, motor speed, verbal fluency, attention and processing speed, and executive functioning. Raw scores were adjusted for age, gender, and education to derive equivalent scores (0–4) for each domain, as well as an overall average equivalent score.

Mental imagery was assessed along two dimensions. Imagery vividness was measured using the visual and kinesthetic subscales of Betts’ Questionnaire upon Mental Imagery (QMI; Sheehan, 1967), adapted into Italian for use with individuals with schizophrenia. Participants rated the vividness of imagined experiences on a 7-point scale, producing total scores ranging from 10 to 70. The ability to maintain and inspect mental images was assessed using two tasks from the Mental Imagery Test (MIT; Di Nuovo et al., 2014), i.e., the Brooks’ “F” Test and the Mental Exploration of a Map task, which together yielded a composite score ranging from 0 to 16.

Psychosocial well-being was measured using the 42-item Italian version of Ryff’s Psychological Well-Being Scales (PWB; Ryff, 1989; Tomyn et al., 2013). The instrument assesses six dimensions of well-being (i.e., Autonomy, Environmental Mastery, Personal Growth, Positive Relations, Purpose in Life, and Self-Acceptance) through items rated on a 6-point Likert scale. Negatively worded items were reverse-scored, and responses were aggregated to obtain a total score ranging from 1 to 252.

### 3.3. The metaphor priming experiment

#### 3.3.1. Materials

##### 3.3.1.1. Metaphorical primes

A set of 40 metaphors (all with an *X is Y* structure, where X is the topic and Y is the vehicle) was selected to be used as metaphorical primes in the experiment. The set included 20 critical metaphor trials and 20 filler metaphors. Critical metaphors were constructed de novo for the present study and involved a vehicle denoting familiar tools or manipulable objects and a topic varying in concreteness (e.g., *Wisdom is a flashlight*). All critical metaphors were rated for familiarity and difficulty by a group of 16 Italian-speaking adults (9 females, Age: *M* = 28.75, *SD* = 7.52; Education: *M* = 18.13, *SD* = 1.31). Participants were asked to judge on a 7-point scale the extent to which each sentence was familiar to them (1 = not familiar, 7 = definitely familiar) and how difficult it was to understand the meaning of the sentence (1 = not difficult, 7 = definitely difficult). On average, critical metaphorical primes were perceived as mostly unfamiliar (*M* = 2.39, *SD* = 1.40, range: 1.25–6.44), yet not too difficult to understand (*M* = 3.08, *SD* = 1.06, range: 1–4.69). Finally, we inspected word-level concreteness of metaphor topics as a proxy for metaphor concreteness, capitalizing on evidence that topic concreteness is strongly correlated with concreteness of the whole metaphorical expression (Bressler et al., 2026). We used GPT-4o (gpt-4o-2024-08-06; OpenAI, 2024) to extract concreteness ratings, in line with prior evidence supporting the alignment of GPT-generated concreteness judgments with human norms (Conde et al., 2026). Following a zero-shot prompting procedure, the model was asked to rate the concreteness of each topic word presented in isolation on a 5-point Likert scale. The final rating was computed as a weighted average of the log probabilities of the three most likely responses, with temperature set to 0, repeating the procedure across five independent runs (Mangiaterra et al., 2025). In the set of critical metaphorical primes, topic concreteness had a mean value of 3.25 (*SD* = 1.30, range: 1.29–4.99).

The remaining 20 metaphors served as filler items and were selected from the Figurative Archive (Bressler et al., 2026), in particular from the dataset in Bambini et al. (2013), which includes norms for familiarity and difficulty.

##### 3.3.1.2. Target words

Each critical metaphorical prime was paired with four different target words, all denoting manipulable objects, corresponding to the four priming conditions: i) semantic, ii) visual, iii) action, or iv) unrelated. The semantic condition was created to test for metaphor-induced semantic priming effects and included target words related to the metaphor vehicle by amodal semantic properties (e.g., meaning association), with no visual or action relatedness (e.g., *lamp* for the prime metaphor *Wisdom is a <u>flashlight</u>*). The visual condition was created to test for metaphor-induced visual priming effects and included target words related to the metaphor vehicle by visual appearance or shape, without strong action or meaning associations (e.g., *microphone* for the prime metaphor *Wisdom is a <u>flashlight</u>*). The action condition was created to test for metaphor-induced action priming effects and included target words related to the metaphor vehicle by manipulability (i.e., the hand-related motor schema required to use the object), with no overt visual or meaning associations (e.g., *remote* for the prime metaphor *Wisdom is a <u>flashlight</u>*). Finally, the unrelated condition included target words without relevant associations with the metaphor vehicle for either amodal or modal properties (e.g., *scissors* for the prime metaphor *Wisdom is a <u>flashlight</u>*). The four types of target words were balanced for log-transformed lexical frequency (*F*(3,76) = 0.87, *p* = .458), extracted from the Corpus and Frequency Lexicon of Written Italian (CoLFIS; Bertinetto et al., 2005), and length as measured in number of syllables (*F*(3,76) = 0.08, *p* = .968) and letters (*F*(3,76) = 1.20, *p* = .316). See Table 5.

**Table 5.** Means and standard deviations for psycholinguistic variables of target words across conditions.

| Measure | Target words |  |  |  |
| --- | --- | --- | --- | --- |
|  | Semantic | Visual | Action | Unrelated |
| Lexical frequency (log) | 3.38 (1.64) | 3.85 (1.59) | 3.02 (1.67) | 3.83 (2.14) |
| Length (in syllables) | 3.25 (1.07) | 3.05 (0.76) | 3.35 (0.88) | 3.25 (0.79) |
| Length (in letters) | 8.20 (2.26) | 7.35 (2.54) | 8.60 (1.82) | 7.95 (1.85) |
| Semantic relatedness | 5.32 (0.81) | 2.19 (1.02) | 2.57 (1.29) | 1.22 (0.13) |
| Visual relatedness | 2.17 (1.24) | 4.78 (1.18) | 2.90 (1.23) | 1.38 (0.51) |
| Action relatedness | 2.67 (1.14) | 3.66 (1.35) | 4.59 (1.09) | 1.57 (0.61) |
| Cosine similarity | 0.35 (0.13) | 0.16 (0.13) | 0.23 (0.10) | 0.07 (0.10) |

*Notes. Relatedness ratings measure the association between metaphor vehicles and each target word (semantic, visual, action, and unrelated) for meaning association, visual properties, and manipulability. Cosine similarity values are computed between metaphorical prime vehicles and each target word.*

To validate our prime-target associations, we conducted a rating task involving 17 Italian-speaking adults (8 females, Age: *M* = 31.18, *SD* = 12.38; Education: *M* = 18.12, *SD* = 1.27), who had not participated in the validation of the metaphorical primes. Participants were presented with word pairs (metaphor vehicles and target words, e.g. *flashlight – lamp*) and were asked to rate the extent to which the two words were related by meaning associations (i.e., semantic relatedness), visual properties (i.e., visual relatedness), and manipulability (i.e., action relatedness) using a 7-point scale (1 = not related, 7 = definitely related). With the exception of the unrelated condition, the mean rating values for all expected associations were higher than 3.5 (Table 5). Specific contrasts between conditions confirmed that semantic targets were rated as more semantically related to the metaphor vehicles than visual (*t*(38) = 10.80, *p* < .001), action (*t*(38) = 8.09, *p* < .001), or unrelated targets (*t*(38) = 22.44, *p* < .001); visual targets were rated as more visually related to the metaphor vehicles than the semantic (*t*(38) = 6.80, *p* < .001), action (*t*(38) = 4.93, *p* < .001), or unrelated targets (*t*(38) = 11.82, *p* < .001); action targets were rated as more manipulation/action related than semantic (*t*(38) = 5.44, *p* < .001), visual (*t*(38) = 2.42, *p* = .020), or unrelated targets (*t*(38) = 10.80, *p* < .001).

To ensure that the prime-target pairs would elicit the expected semantic priming effects, we also computed the semantic similarity between metaphorical prime vehicles and the associated target words as an additional check, extracted using *LSAfun* R package (Günther et al., 2015) with a semantic space built on ItWac corpus (Baroni et al., 2009): the cosine similarity between the metaphorical prime vehicles and the targets was significantly lower for unrelated compared to semantic (*t*(36) = 7.32, *p* < .001), visual (*t*(36) = 2.36, *p* = .024), action targets (*t*(33) = 4.76, *p* < .001), while it was significantly higher for semantic compared to visual (*t*(36) = 4.47, *p* < .001) and action (*t*(33) = 2.83, *p* = .007) targets (Table 5).

#### 3.3.2. Procedure

The priming experiment was implemented as a metaphor-primed lexical decision task. Each participant was randomly assigned to a single ISI group (400, 1000, and 1400 ms) and was presented with 120 trials (80 critical and 40 filler trials), divided into four blocks with a short break between blocks. Items and blocks were randomized. After a fixation cross (500 ms), the metaphorical prime appeared on the screen for 1500 ms followed by a variable ISI. Afterwards, the target word appeared and lasted on the screen for 2000 ms, during which the participants were asked to decide whether the target was an existing Italian word or not by pressing one of two buttons on a Cedrus® RB-540 response pad. In critical trials, the target was always a real word, while filler trials contained a pseudo-word (e.g., *matolca*). The button order was counterbalanced across participants to control for handedness. Following the response window, the next trial automatically began after a 500 ms blank screen. Before the experimental sessions, participants completed 10 practice trials. The trial structure and the rationale of the procedure is exemplified in Figure 1. The experiment was implemented on PsychoPy© software (Peirce et al., 2019).

After the metaphor priming experiment, an offline multiple-choice metaphor comprehension task was administered to ensure that participants were able to understand the prime metaphors, modelled after standardized tests including multiple-choice tasks to evaluate metaphor comprehension (Arcara & Bambini, 2016; Bischetti et al., 2025). Participants were presented with the metaphors used as primes in critical trials during the experiment (one item at a time) on a laptop monitor. Metaphors were presented alongside three potential interpretations (correct, incorrect concrete, and incorrect unrelated interpretations) and were read aloud by the experimenter to minimize memory loading. Participants were asked to select the correct interpretation among the three, and a binomial score (1 = correct, 0 = incorrect) was assigned for each item (score range: 0–20).

### 3.4. Statistical Analysis

Preliminarily, we run a series of independent-samples *t*-tests to test whether patients and controls differed in the assessment measures (i.e., lexical and semantic skills, pragmatic abilities, neurocognition, psychosocial well-being, and mental imagery). The *t*-tests’ normality assumption was checked by visual inspection of the distribution of the dependent variables, while the homogeneity of variance assumption (homoskedasticity) was checked with tests of variance (using the *var.test()* function in R). Welch’s independent sample *t*-test was used in the case of heteroskedasticity. All *p-*values were adjusted for False Discovery Rate (FDR; Benjamini & Hochberg, 1995).

Then, we tested for the presence of altered semantic and sensory-motor activation in the metaphor priming experiment. Semantic priming effects were tested using the unrelated target as the baseline (Unrelated RTs – Semantic RTs), while for visual and action priming effects we used the semantic target as baseline (Semantic RTs – Visual/Action RTs), as we reasoned that vision and action priming should be compared to an amodal baseline, in line with neuroimaging literature testing whether of multimodal representations explain more variance than unimodal models (Tong et al., 2022). We fitted a Linear Mixed-effects Model on participants’ *z*-transformed metaphor-primed lexical decision latencies using *lme4* and *lmerTest* packages (Bates et al., 2015; Kuznetsova et al., 2017), including Condition (within subjects, four levels: Visual, Action, Semantic, and Unrelated) in interaction with Group (between subjects, two levels: Patients and Controls, sum-contrast coded as Controls = 0.5, Schizophrenia = –0.5) as fixed factors nested within ISI (between subjects, three levels: 400, 1000, and 1400 ms). We constructed the contrast coding matrix for Condition in a way that would allow us to test for semantic, visual, and action priming effects in the same model (Semantic Priming: Unrelated – Semantic; Visual Priming: Semantic – Visual; Action Priming: Semantic – Action). The optimal random structure was always determined in a stepwise fashion using the *buildmer* package (Voeten, 2023), starting from a maximal model including all possible fixed and random effects. The final random structure included by-subjects and by-items random intercepts only, as more complex structures did not lead to model convergence. Prior to running any models on latencies, RTs were pre-processed and cleaned by excluding all participants with < 70% of correct answers in the metaphor-primed lexical decision task (*n* = 2 participants from the schizophrenia group and *n* = 2 participants from the control group were removed). We also trimmed trials that were: i) incorrect, ii) faster than 250 ms, and iii) exceeding |2.5| standard deviations from participants’ means across conditions (4.34% and 6.81% deleted trials for controls and patients, respectively). Finally, only responses for which the full condition quadruplet (i.e., unrelated, semantic, action, and visual) was available were included in the analysis.

Afterwards, we tested whether semantic or sensory-motor activations were predictive of accuracy in the offline metaphor comprehension task, and whether they were associated with any measure collected during the offline behavioral assessment. To do so, we first inspected correlograms with patterns of correlations in each relevant ISI, computed for the schizophrenia and control groups separately, using Spearman’s correlations. We then tested whether priming effects predicted metaphor comprehension scores, controlling for the role of known predictors of metaphor comprehension available in both groups (i.e., lexical-semantic and neurocognitive skills), by fitting a multiple regression model, with the total score of the metaphor comprehension task as the dependent variable nested across relevant inter-stimulus intervals and groups, and priming scores (aggregated by subjects) as independent variables, with WAIS-R Vocabulary subtask and the BACS average equivalent as covariates. Before running Spearman’s correlations and the multiple regression model, we checked that the different ISI subgroups in the schizophrenia and control samples were not significantly different for individual difference variables. A series of One-way ANOVA tests showed that the participants from the schizophrenia group assigned to the different ISI subgroups did not differ in any individual difference variable (*F*s < 0.64, *p*s > .529), namely demographic (i.e., age and education), clinical (i.e., illness onset and duration, psychopathology severity, and mean chlorpromazine equivalent dose), and cognitive measure (i.e., lexical-semantic, neurocognitive, and imagistic skills), except for the PANSS General score, which was lower for participants assigned to the ISI 1000 subgroup (*F*(2,59) = 4.21, *p* = .020; pairwise post-hoc comparisons: ISI 400 > ISI 1000; ISI 1000 = ISI 1400; ISI 400 = ISI 1400). Similarly, the participants from the control group did not differ across ISI subgroups for any demographic and cognitive measure (*F*s < 2.29, *p*s > .109).

Finally, we inspected whether semantic and sensory-motor priming effects would be predicted by item-level properties of the metaphorical primes, in particular the familiarity of the metaphor and the topic concreteness. To do so, we computed Pearson’s correlations between priming scores aggregated at item level and familiarity and topic concreteness ratings, limiting the exploration only to conditions where we found significant priming effects.

The analysis was performed using RStudio (R Core Team, 2024), version 2023.12.0+369.

## 4. Data availability

The materials and data associated with this study are publicly available in the Zenodo repository (https://doi.org/10.5281/zenodo.21828307). The materials include metaphorical primes and target words, along with their properties (i.e., psycholinguistic variables and values of de novo collected ratings). The data include the performance of participants with schizophrenia and healthy controls in the metaphor priming experiment and in the metaphor comprehension task.

## 6. Author contribution (CRediT)

V.B.: Conceptualization; Methodology; Funding Acquisition; Supervision; Project Administration; Writing – Original Draft; F.F.: Methodology; Software; Investigation; Data Curation; Formal Analysis; Visualization; Writing – Original Draft; C.P.: Investigation; Data Curation; L.B.: Methodology; Validation; V.M.: Investigation; G.M.: Software; C.B.: Investigation; L.V.: Investigation; Data Curation; G.A.: Investigation; Data Curation; M.Be.: Investigation; Data Curation; M.Bu: Investigation; Data Curation; J.S.: Investigation; Data Curation; F.M.: Investigation; Data Curation; M.S.: Investigation; Data Curation; F.C.: Investigation; Data Curation; R.C.: Resources; Writing – Review & Editing; M.B.: Conceptualization; Resources; Supervision; Project Administration; Writing – Review & Editing.

## Data Availability

The materials and data associated with this study are publicly available in the Zenodo repository (https://doi.org/10.5281/zenodo.21828307).

